# Evaluation of a Virtual Reality-Based Eye Tracker for Neuro-Ophthalmic Assessment: A Feasibility, Reliability and Reproducibility Study

**DOI:** 10.1101/2025.06.18.25329788

**Authors:** Irem Karaer, Ha-Jun Yoon, Runfeng Ma, Reenette Savant, Vanessa Rodwell, Riddhi Shenoy, Zhanhan Tu, Qadeer Arshad, Elizabeta B Mukaetova-Ladinska, Mervyn G Thomas

## Abstract

**Background/Objectives:** Virtual Reality (VR) eye trackers offer portable, objective tools for neuro-ophthalmic testing. This study evaluated the feasibility, reproducibility and reliability of a VR eye tracker (BulbiCAM) compared to wearable eye-tracking glasses (PupilLabs Neon glasses), highlighting its potential clinical utility and feasibility.

**Subjects/Methods:** A prospective study involving 39 healthy participants (mean age⍰±⍰SD = 30.0 ± 9.5 years) assessed inter-visit reproducibility of BulbiCAM tests across two visits. Pupillary light reflex tests were conducted with both BulbiCAM and PupilLabs Neon, enabling paired assessments. Reproducibility was analysed using intra-class correlation coefficients (ICC), reliability via Bland-Altman analysis, and participant experience through a survey evaluating test comfort and usability.

**Results:** Participants’ feedback (n=27) highlighted high acceptability for BulbiCAM: 89% found the test comfortable, 92.6% felt the testing duration was appropriate, and 81.5% reported no eye strain or fatigue. Inter-visit reproducibility of Bulbicam tests showed high reproducibility for pursuit and pupil tests (ICC= 0.88-0.76), while saccadic tasks showed lower reproducibility (best ICC at 0.62). Paired assessments between devices showed close agreement for key pupillometer metrics: baseline diameter (bias: -0.48 ± 0.47 mm), peak constriction diameter (bias: -0.56 ± 0.36 mm), constriction velocity (bias: 0.22 ± 0.58 mm/s), and duration of constriction (bias: -0.052 ± 0.15 s).

**Conclusions:** This study highlights the clinical feasibility of BulbiCAM, with high patient acceptability and reproducibility for pursuit and pupil tests. Paired assessments confirmed its accuracy for key pupillometric parameters, validating its reliability for clinical and research use.

## Introduction

Eye-tracking devices are widely utilized in both neuro-ophthalmic research and clinical settings, providing valuable insights into ocular and neurological function (1–3). In recent years, Virtual Reality (VR) headset eye-tracking devices have gained increasing popularity as portable and efficient alternatives to conventional eye-tracking systems (4). By integrating advanced eye-tracking capabilities within VR headsets, these devices offer enhanced flexibility and convenience (5). Unlike conventional setups with separate displays, projectors, and computers, these all-in-one systems offer a space-saving solution for clinics while providing accurate eye-tracking data.

Both subjective and objective methods are employed to evaluate eye movements and pupillary light reflexes. However, there is a growing need for clinically adaptable eye trackers utilizing objective methods (6). Particularly, VR headset eye-tracking devices equipped with automated software, offer significant advantages by providing precise and standardized measurements, ensuring accuracy and reliability (7,8). BulbiCAM is an example of a VR headset with eye tracking capabilities aimed at obtaining objective metrics for oculomotor behaviour and pupillometer; thus having the potential for assessing a range of neuro-ophthalmic diseases and dysfunctions (9).

VR eye tracking systems are under-explored compared to traditional systems (10) and to date, there are no reproducibility/reliability studies using VR headset-based eye trackers. Therefore, in this study, we aim to assess the reproducibility of BulbiCAM tests and reliability of BulbiCAM compared to other wearable eye tracker with pupillometry such as PupilLabs Neon glasses, offering new insights into their potential clinical applicability.

## Materials (Subjects) and Methods

### Participants

Healthy participants (n=39) were recruited for the study and successfully completed both visits. Healthy subjects were defined as participants who had no neurological diseases or clinical signs of ocular motility dysfunction. The demographic characteristics of the participants are presented in Table 1. Their ages ranged from 21 to 59 years, and the male-to-female ratio was 18:22. Refractive errors ranged from -8.00D to +5.50D. All subjects achieved a best-corrected visual acuity (BCVA) of 0.20 logMAR or better. Colour vision, assessed with the Ishihara pseudoisochromatic plates, was normal (17/17) in most participants. However, one individual recorded scores of 5/17 in the right eye and 6/17 in the left eye.

**Table 1:** Demographic information of participants.

| **Variable** |  |
| --- | --- |
| **Age, Mean ± SD** | **30.2±9.7** |
| **Sex, No. (%)** |  |
| Female | 21(54) |
| Male | 18(46) |
| **Ethnicity, No. (%)** |  |
| White-British | 2 (5) |
| White-Other | 8 (18) |
| Asian-Oriental | 21 (54) |
| Asian-Indian Subcontinent | 7 (18) |
| Afro-Caribbean | 1 (2.5) |
| Other | 1(2.5) |
| **Visual Acuity (corrected, logMAR), Mean ± SD** |  |
| Left eye | -0.01±0.06 |
| Right eye | -0.02± 0.07 |
| **Colour Vision** |  |
| Normal No. (%) | 38 (97.4) |

None of the participants had a history of neurological disorders. Ophthalmological histories were largely unremarkable, except in five individuals: three had corneal refractive surgery, one had previously undergone strabismus surgery and also had colour blindness, and one had lattice degeneration and posterior vitreous detachment in both eyes.

Written informed consent was obtained from each participant.

### Study Design

39 participants completed two testing sessions, scheduled at similar times of the day. At baseline, all participants underwent best-corrected visual acuity assessments (ETDRS chart at 4m) and Ishihara tests for colour vision screening.

Figure 1 illustrates the participants’ flow through the study and the order of testing performed. Further details regarding the stimulus parameters used in BulbiCAM are provided below.

**Figure 1:**
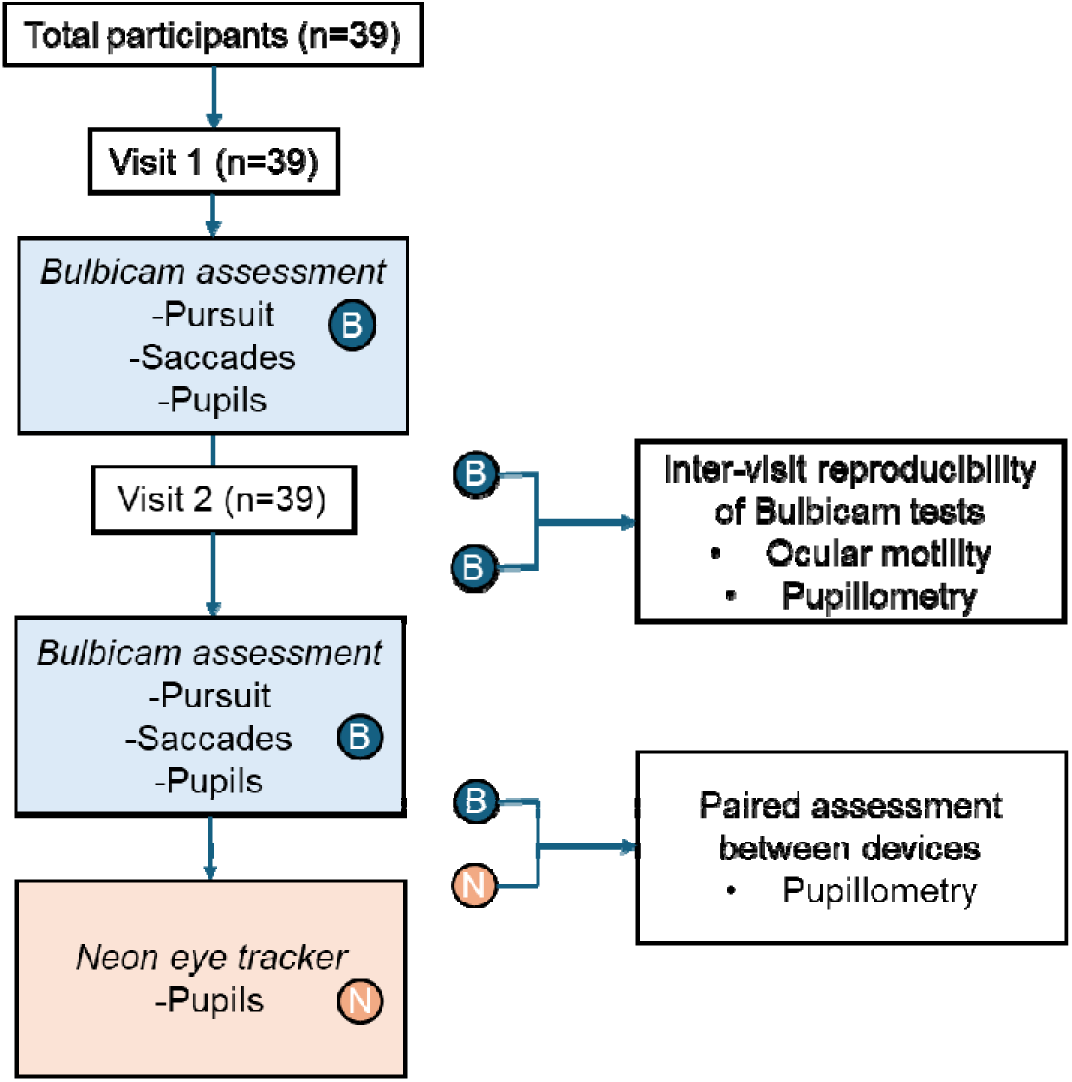
Flowchart explaining the study design. B: Bulbicam, N: PupilLabs Neon eye tracking glasses

In the segment of the study involving Neon glasses, participants wore the glasses while tracking stimuli displayed on the BulbiCAM screen. During these measurements, participants’ heads were positioned slightly behind the standard posture to accommodate the glasses and ensure accurate data capture.

### Participant Experience

Participants were asked to complete a questionnaire after completing two visits. The questionnaire scored (1) Comfort, (2) Length of the examination, (3) Brightness, (4) Double vision, (5) Grading the fatigue/ eye strain specifically for Bulbicam on a 5-point Likert scale as: 1 = Strongly Disagree, 2 = Disagree, 3 = Neutral, 4 = Agree, 5 = Strongly Agree (Figure 3).

### BulbiCAM

BulbiCAM (BulbiTech AS, Oslo, Norway) is a CE-certified and FDA-registered eye-tracking device that uses AI-integrated software to assess the data obtained by cameras. It accommodates an interpupillary distance of 49mm to 75mm. The display mode hardware utilised is the Sharp LS055R1SX04, which consists of 5.5” LCD panel (2560×1440 pixels @60 fps; 1920×1080 pixels @60 fps), MIPI board, White 60-pin flex cable, HDMI-MIPI board (based on TC358870XBG M silicon). The camera is configured to capture two different types of images alternately, Dark Pupil (DP) and Bright Pupil (BP). If the initial frame is BP, the next one will be DP, followed by BP again, and so on. The camera operates at 400 frames per second (FPS), capturing 200 DP and 200 BP images per second, resulting in a total of 400 pupil images per second. The stimulus synchronization typically has an accuracy of ±18 ms, with a worst-case deviation of ±38 ms (9).

Experimental set-up is shown on Figure 2A, and Figure 2B shows real time recording screen.

**Figure 2:**
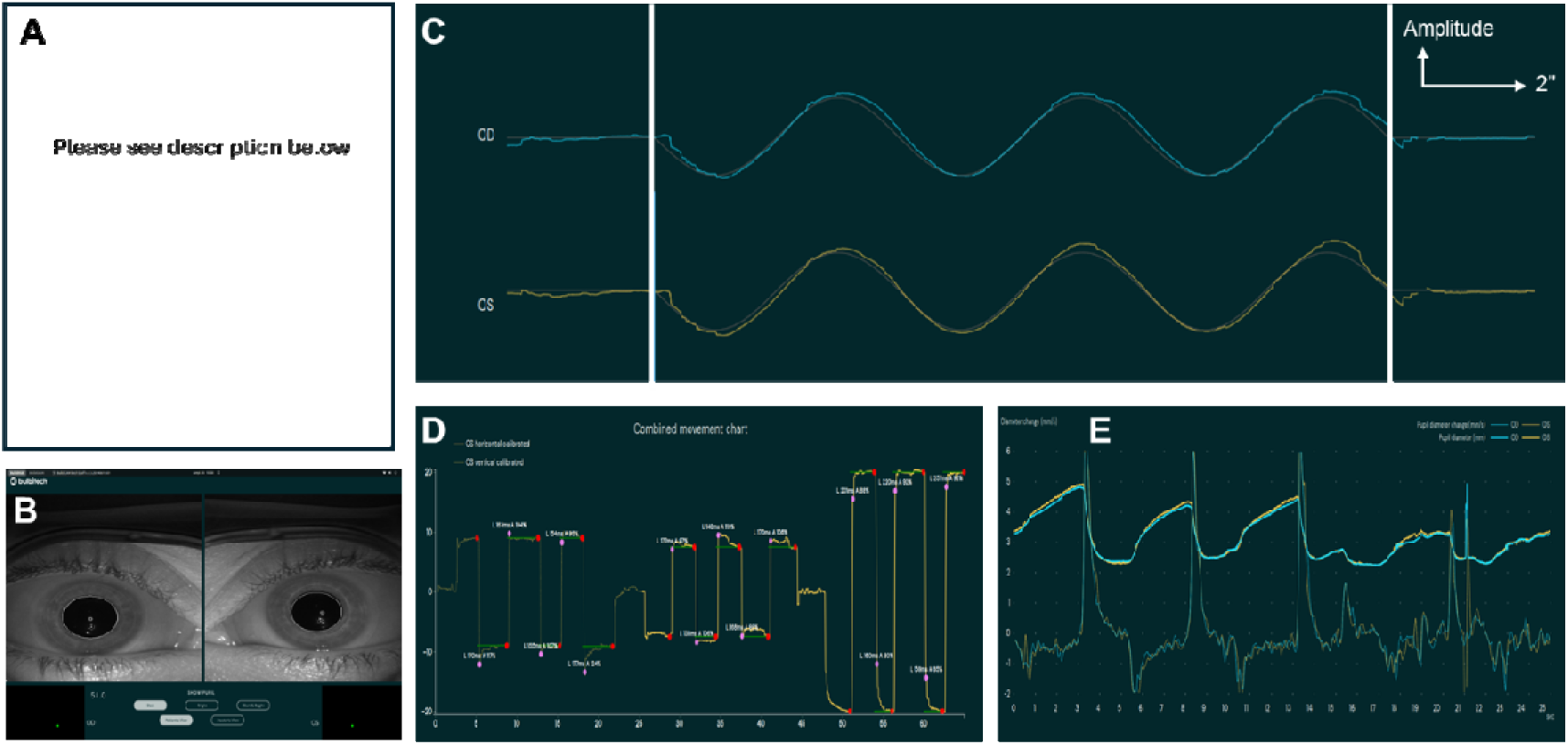
(A) BulbiCAM eye tracking device set up and a participant being assessed (**please contact the corresponding author to request access to Figure2A**) (B) Bulbicam screen during assessment (C) original pursuit test data. White lines show the starting and ending of the stimulus. Vertical arrow is a scale of wave amplitude and horizontal arrow represents two seconds (D) original saccades test data (E) original pupil test dataFigure 3: Patient Experience Across Different Variables. Bar charts illustrating the frequency of responses for various patient experience factors: (A) Comfort during the procedure, (B) Appropriateness of the examination length, (C) Brightness levels used during the examination, (D) Incidence of double vision, and (E) Eye strain or fatigue experienced during or after the examination.

### PupilLabs Neon glasses

PupilLabs Neon Glasses (Pupil Labs GmbH, Berlin, Germany) are eye-tracking glasses equipped with scene cameras in front of the glasses and two eye cameras with 850 nm infrared illuminator LEDs. The device is connected to the companion device (Android phone) which enables the glasses to run and process the data collection. Pupil Cloud website is the recommended tool for analysing the data and processing the recordings. PupilLabs Neon glasses enables researchers to collect gaze, saccades, pupil diameters data alongside real-world front camera footage. The camera operates at a frequency of 200 frames per second (FPS) (11).

### Data Set

Pursuits, saccades and pupillary light reflex test values were obtained by BulbiCAM. The smooth pursuit test evaluates how well the eye tracks a visual target smoothly. The gain values were provided for pursuit tests in response to 10 degree 0.5 Hz frequency stimuli. Gain, which measures tracking accuracy , is defined as the ratio of the stimulus’s velocity to the eye’s movement velocity. Perfect tracking is indicated by a gain value of 100%, but under-tracking is suggested by values less than 100%. Through examination of this variable at different stimulus frequencies, the accuracy and effectiveness of the smooth pursuit system can be evaluated in various scenarios (9,12). Stimuli and tracking recordings of both right and left eye is shown in Figure 2C.

Saccades are rapid eye movements that shift gaze between different targets. During a visual search task, the saccade test delivers both horizontal and vertical stimuli. Peak velocity, latency, and accuracy parameters are provided to assess saccadic eye movements. Vertical 9 degree and horizontal 20 degree saccade parameters were assessed using BulbiCAM’s standard test protocol. Figure 2D shows examples of horizontal and vertical saccades.

Pupil baseline-peak constriction diameters and constriction velocity values were obtained for pupillometry testing. Data from only one eye was analysed to assess reproducibility and reliability of pupillometry testing, since all the participants were healthy and we expected a high degree of correlation between the two eyes. The diagram in Figure 2E shows the pupil diameter increasing and decreasing through the trials. In addition to Bulbicam, the same variables for pupil test were obtained from PupilLabs Neon glasses Pupil Cloud platform where gaze and pupil diameter data is available.

### Statistical Analysis

A sample size calculation was performed based on an expected intraclass correlation coefficient (ICC) of 0.675, derived from saccadic gain reliability (ICCs ranged from 0.52 – 0.83) data in healthy controls reported in a Huntington’s disease study (13). Assuming a null hypothesis ICC of 0.50, a significance level of 0.05, and 80% power, the required sample size was estimated to be 27 participants. To account for an anticipated 10% dropout rate, the final adjusted sample size was determined to be 31 participants.

The ICC was used to assess inter-visit reproducibility of BulbiCAM tests (14). At the end of the two visits, data from n=39 participants was analysed for the pursuit test, n=37 for the vertical 9 degree saccadic test, n=35 for the horizontal 20 degree saccadic test, and n=35 for the pupil test, respectively. For pursuits, gain values were analysed in response to 10 degree 0.5 Hz frequency stimulus. For saccades, peak velocity, latency and accuracy values of vertical 9 degree and horizontal 20 degree were analysed. For the pupil test; baseline pupil diameter, peak constriction diameter and constriction velocity were analysed. Test reproducibility is categorized as excellent when the ICC exceeds 0.90, good when it falls between 0.75 and 0.90, moderate for values ranging from 0.50 to 0.75, and poor when it is below 0.50 (15).

We obtained paired pupillometry data from PupilLabs Neon glasses in n=23 participants and used Bland-Altman test to evaluate the agreement in pupillary light reflex measurements between BulbiCAM and PupilLabs Neon glasses (16).

Bland-Altman analysis was performed using GraphPad Prism, and ICC analysis using R.

### Ethical approval

This study received ethical approval from the East Midlands – Nottingham 2 Research Ethics Committee (REC) (REC reference: 21/EM/0017, protocol version: 0772, and the UK Health Research Authority (HRA) approval (IRAS ID: 279309).

## Results

### Participant Experience

A total of n=27 participants provided feedback on their BulbiCAM examination experience using a 5-point Likert scale, achieving an excellent response rate, with majority reporting positive experiences. Specifically, 89.0% found the test comfortable, 92.6% considered the examination length appropriate, and 92.5% were satisfied with the brightness levels. Furthermore, 62.9% reported no occurrence of double vision, and 81.5% indicated they experienced no eye strain or fatigue (Figure 3).

**Figure 3:**
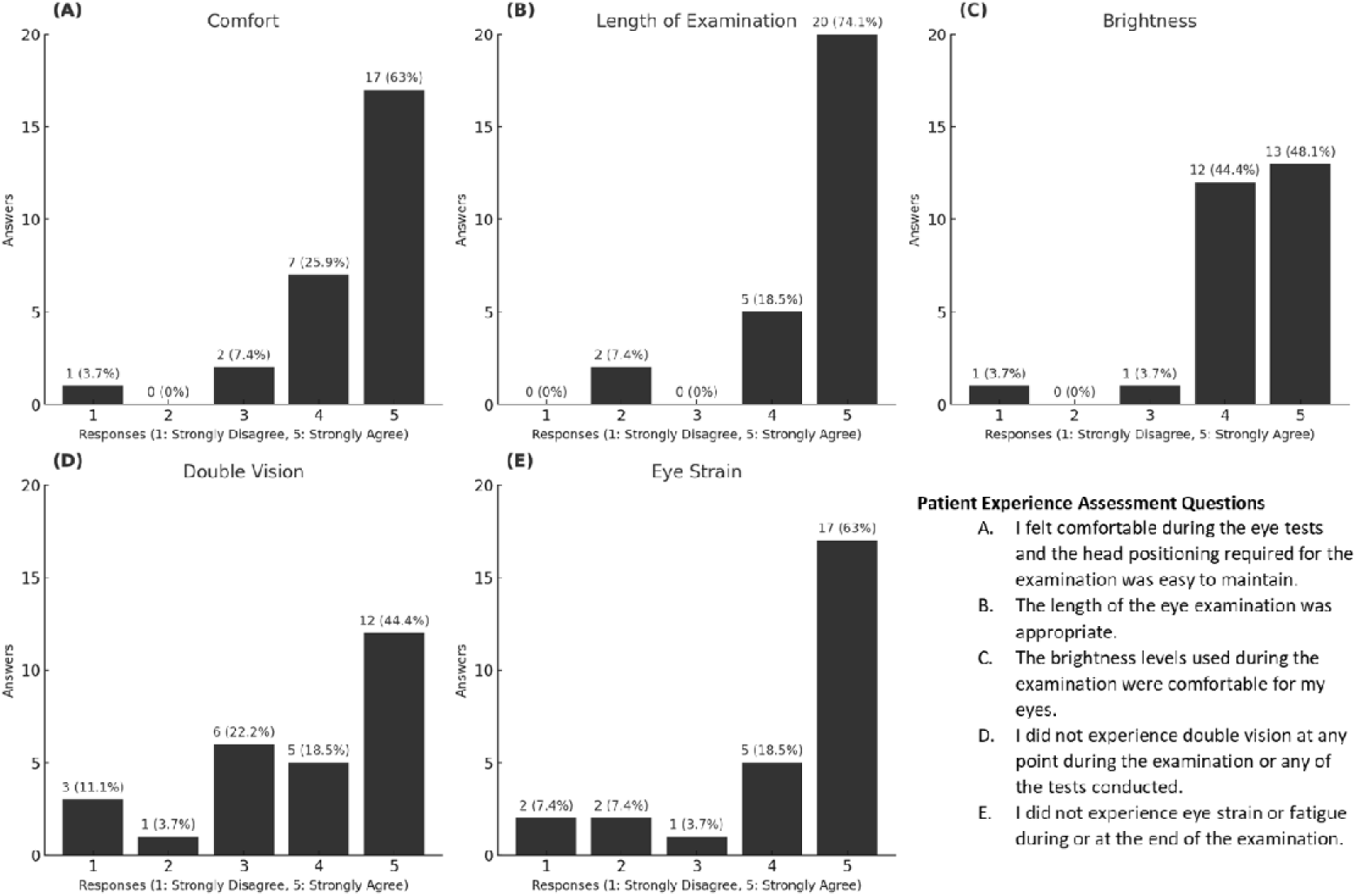
Patient Experience Across Different Variables. Bar charts illustrating the frequency of responses for various patient experience factors: (A) Comfort during the procedure, (B) Appropriateness of the examination length, (C) Brightness levels used during the examination, (D) Incidence of double vision, and (E) Eye strain or fatigue experienced during or after the examination.

The pursuit test demonstrated high reproducibility, with an ICC value of 0.81 (Figure 4A). For the vertical 9 degree saccades test, the ICC values for latency, peak velocity, and accuracy were 0.62, 0.50, and 0.49, respectively. Similarly, for the horizontal 20 degree saccades test, the ICC values for these metrics were 0.52, 0.46, and 0.61, respectively (Figure 4A).

**Figure 4.**
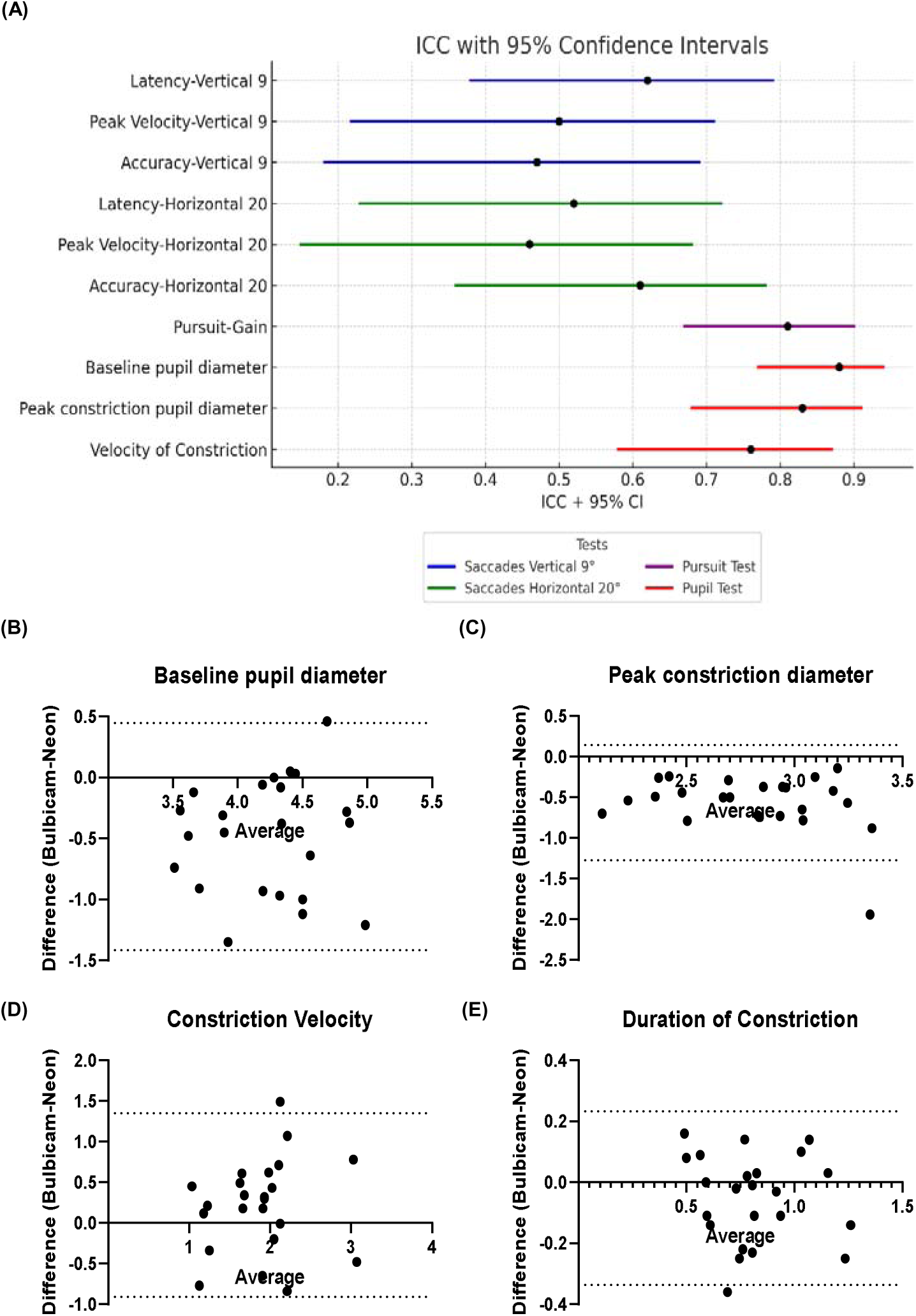
Intra-Class Correlation (ICC) Analysis and Bland-Altman Plots: (A) ICC values with 95% confidence intervals for saccades, pursuit and pupil tests. Colours represent different test groups: vertical 9°(degree) saccadic task (blue), horizontal 20°(degree) saccadic task (green), pursuit test (purple), and pupil tests (red). (B-E) Bland-Altman plots for the pupil test metrics for pupil diameter (baseline and peak constriction), constriction velocity (mm/s) and duration of constriction. Average represents the bias between two measurements, while dashed lines indicate the 95% limits of agreement (LoA).

In the pupil test, high reproducibility was observed for the baseline pupil diameter (ICC: 0.88), for the peak constriction diameter (ICC: 0.83), as well as for the constriction velocity (ICC: 0.76) (Figure 4A).

These results indicate varying levels of reproducibility across the tested parameters, with particularly strong consistency in the pursuit and pupil test metrics.

### Paired Assessment Between Devices

The Bland-Altman plots illustrate the agreement between BulbiCAM and PupilLabs Neon glasses measurements across various pupil test parameters: Paired assessment between devices showed close agreement for key pupillometer metrics: baseline diameter (bias: -0.48 ± 0.47 mm LoA:-1.4 to 0.45), peak constriction diameter (bias: -0.56 ± 0.36 mm LoA: -1.3 to 0.15), constriction velocity (bias: 0.22 ± 0.58 mm/s LoA: -0.9 to 1.3), and duration of constriction (bias: -0.052 ± 0.15 s LoA: -0.34 to 0.23). Most data points fell within the limits of agreement (LoA), demonstrating agreement between systems (Figure 4B, 4C, 4D, 4E). These results suggest good agreement between the two systems, with minor, consistent biases across static and dynamic pupil measurement parameters.

## Discussion

This study evaluated the feasibility of using the BulbiCAM device in a clinical setting, focusing on the reproducibility of pursuit, saccade, and pupillometry tests. In addition, a paired assessment between BulbiCAM and the PupilLabs Neon eye-tracking glasses examined the agreement between devices. Overall, participant feedback indicated a high level of acceptability, with favourable ratings for comfort, examination length, brightness, and minimal reports of double vision or eye strain. Smooth pursuit and pupillometry tests demonstrated strong reproducibility, while saccade tests showed more variable results. These findings support the potential everyday clinical applicability of BulbiCAM as a feasible and well-tolerated eye-tracking solution.

### Inter-visit Reproducibility of BulbiCAM Tests

The pursuit test showed good reproducibility at 10-degree, 0.5 Hz test frequency. The reproducibility of the BulbiCAM was assessed using the gain value of the pursuit test at a stimulus frequency of 0.5 Hz, as pursuit gain is known to decrease at frequencies above 0.5 Hz (17). Although previous studies have shown that gain values at 0.25 Hz are higher than those at 0.5 Hz in children, with performance declining as target velocity increases, our findings confirm the high reproducibility of the pursuit test at 0.5 Hz. This suggests that 0.5 Hz is an ideal parameter for reproducibility assessments (18,19).

Inter-visit reproducibility for pupillary light reaction tests showed good reproducibility for baseline-peak constriction diameters and constriction velocity parameters. In contrast, the latency, peak velocity, and accuracy parameters of the vertical 9 and horizontal 20 saccade tests showed only moderate to poor reproducibility. This discrepancy in reproducibility between the saccadic and pupillometry tests can be attributed to fundamental differences in the physiological mechanisms underlying these tasks. Pupillary light reflexes are primarily mediated by the autonomic nervous system, which is relatively stable and less influenced by higher-order cognitive processes.

In contrast, saccadic eye movements are complex, involving the integration of sensory, cognitive, and motor systems. Factors such as attention, learning effects, and decision-making are known to influence saccadic performance, potentially introducing variability between visits (20–22). Although previous studies have reported high reproducibility for saccade tests under controlled conditions (15), the moderate to poor reproducibility observed in our study may reflect individual differences in attention and engagement across visits. Notably, the pursuit tasks exhibited less variability, despite also being influenced by attention and engagement. This difference may be attributed to the pursuit system’s reliance on continuous, reflexive tracking mechanisms, which are less susceptible to transient fluctuations in attention, cognitive engagement, or physiological states compared to saccadic movements. Additionally, the study design, which required two separate visits, could have introduced variability due to learning effects or differences in participant familiarity with the task.

The use of a VR-based testing system, such as the BulbiCAM, might also play a role. While effective for certain measures, it may not be optimized for capturing the fine-grained dynamics of saccadic eye movements with high reproducibility. Future studies could explore modifications to the testing protocol or alternative setups to improve the reliability of saccade measurements using VR-based systems.

### Paired Assessment Between Devices

This study demonstrates agreement between devices in pupillometry test variables, indicating reliable measurements across the observed parameters. BulbiCAM consistently measured lower pupil diameters for both baseline and peak constriction states, compared to PupilLabs Neon glasses. These differences can be attributed to variations in participants’ head positioning during the assessments. During Neon glasses testing, participants’ heads were positioned slightly back, leading to variable brightness levels. In contrast, BulbiCAM assessments ensured more consistent head positioning, resulting in brighter light exposure and consequently smaller pupil diameters, particularly at the peak constriction phase. The baseline pupil diameters were also measured lower with BulbiCAM, likely due to similar factors. Although the variable is termed as “baseline” diameter, repeated exposure to bright light stimuli during the trials (as mentioned previously on methodology section) is likely to influence the baseline diameters. This effect, known as the post-illumination pupil response, reflects the delay in returning to baseline dilation after exposure to a stimulus, which can take several seconds (23). Consequently, differences in stimulus illumination levels contributed to the variation in baseline pupil diameter in this study.

Discrepancies in baseline measurements between BulbiCAM and PupilLabs Neon glasses are primarily attributed to variations in head positioning and light illuminance, with a previously published comparison study similarly highlighting the effect of target illuminance variability on different measurement methods (24). Despite these factors, the observed differences in baseline and peak constriction pupil diameters still fall within acceptable limits in our study. Schmitz et al (2003) previously compared three different methods of pupil diameter measurement and suggested that deviations of up to 1 mm may be acceptable if clinically insignificant (25). Considering the findings of this study alongside previous research and the potential effects of varying stimulus illuminance, the agreement between BulbiCAM and PupilLabs Neon glasses for pupil diameter measurements remains robust and clinically applicable.

The BulbiCAM system measures a higher velocity of pupil constriction compared to Neon glasses. Pupil velocity is determined by identifying a consistent constriction or dilation phase and calculating the change in pupil diameter or area divided by the phase duration (12). Velocity, defined as the change in pupil diameter per second for this test, is greater in BulbiCAM because the diameter change (absolute value of baseline-peak constriction difference) is larger, and the time to reach peak constriction is faster. Neon glasses recorded a longer time to constriction, likely due to lower brightness levels during the assessment, which may have caused a slower and more gradual pupil constriction compared to the sharper response observed with BulbiCAM. Therefore, while BulbiCAM shows higher velocity measurements, considering the diameter change and time to constriction variables used to calculate velocity, BulbiCAM’s velocity measurement is still acceptable and reliable for pupil testing.

These results reinforce the reliability of both devices for pupillometry assessments while acknowledging the influence of experimental conditions on measurement outcomes.

### Limitations

As a limiting factor of this study, reliability was assessed solely for the paired comparison of the pupil test between BulbiCAM and PupilLabs Neon glasses. No evaluations involving other devices or tests were conducted, as replicating the exact stimulus was not feasible with a traditional infrared eye tracker.

Additionally, the recent updates to the device software, require further testing to assess their impact on reproducibility and reliability.

## Conclusion

This study demonstrates the feasibility, reproducibility and reliability of the BulbiCAM as a tool for both clinical and research applications. Patient feedback on the BulbiCAM examination was highly positive. Inter-visit reproducibility was high for pursuit and pupil tests, supporting the device’s consistency. While saccades showed poor to moderate reproducibility, this highlights areas for potential refinement and investigating further. The paired assessment between devices further validated the accuracy of key pupillometric parameter. These results underscore the potential of BulbiCAM as a reliable and patient-friendly device for oculomotor and pupillometric evaluations in both clinical and research settings.

The findings of this study provide a foundation for future research into the application of VR-based eye-tracking systems across various disorders and contexts. In future, we foresee portable VR based headset eye tracking devices playing a significant role in meeting the increasing need for imaging across diverse settings, confirming the point-of-care approach.

## Data Availability

All data produced in the present study are available upon reasonable request to the authors

## Acknowledgements

We thank Vincent Dwianta and Gurtek Samra for their insight on PupilLabs Neon glasses setup. We thank Dennis Hens and the BulbiCAM technical team for troubleshooting technical challenges as required during the study.

## Author contributions

MT, IK: study design; MT: supervision of the project; IK, RM: data collection; MT, IK, HY: data extraction analysis; IK, RS: drafting the original manuscript; MT, EML, QA: funding acquisition; MT, IK, EML, QA, VR, HY, RS, RM, RS, ZT: critical review of the manuscript.

All authors have given their approval for the published version of the manuscript.

## Summary

### What was known before?

- Eye-tracking devices are essential in neuro-ophthalmic research and clinical practice, offering key insights into ocular and neurological function.
- VR headset-based eye trackers offer objective assessment methods in a portable, all-in-one compact system.
- VR eye tracking systems are under-explored compared to traditional systems and no reproducibility or reliability studies have been conducted on VR headset-based eye trackers to date.

### What this study adds?

- Participant feedback highlighted high acceptability; while paired assessment between devices (BulbiCAM and PupilLabs Neon eye tracking glasses) showed strong agreement confirming BulbiCAM’s reliability.
- VR headset eye tracking device Bulbicam demonstrated high reproducibility for pursuit and pupil tests; while saccade tests had lower reproducibility outcomes.
- In future, portable VR headsets could play a key role in expanding point-of-care assessments in various settings.

## References

1. Oyama A, Takeda S, Ito Y, Nakajima T, Takami Y, Takeya Y, et al. Novel Method for Rapid Assessment of Cognitive Impairment Using High-Performance Eye-Tracking Technology. Sci Rep 2019 Sep 10;9(1):12932–x.

2. Kim M, Lee J, Lee SY, Ha M, Park I, Jang J, et al. Development of an eye-tracking system based on a deep learning model to assess executive function in patients with mental illnesses. Sci Rep 2024 Aug 6;14(1):18186–2.

3. Leigh RJ, Kennard C. Using saccades as a research tool in the clinical neurosciences. Brain 2004;127(3):460–477.

4. Xu Y, Zhang C, Pan B, Yuan Q, Zhang X. A portable and efficient dementia screening tool using eye tracking machine learning and virtual reality. NPJ Digit Med 2024 Aug 22;7(1):219–5.

5. Moreno-Arjonilla J, López-Ruiz A, Jiménez-Pérez JR, Callejas-Aguilera JE, Jurado JM. Eye-tracking on virtual reality: a survey. Virtual Reality 2024 -02-05;28(1).

6. Hirota M, Kato K, Fukushima M, Ikeda Y, Hayashi T, Mizota A. Analysis of smooth pursuit eye movements in a clinical context by tracking the target and eyes. Sci Rep 2022 May 19;12(1):8501–6.

7. THE POWER OF SIGHT: USING EYE TRACKING TO ASSESS LEARNING EXPERIENCE (LX) IN VIRTUAL REALITY ENVIRONMENTS. 11th International Technology, Education and Development Conference; 6-8 March, 2017; INTED2017 Proceedings: 2017.

8. Barkevich K, Bailey R, Diaz GJ. Using Deep Learning to Increase Eye-Tracking Robustness, Accuracy, and Precision in Virtual Reality. Proc ACM Comput Graph Interact Tech 2024 -05-17;7(2):1.

9. Bulbitech user manual. 2023; Available at: https://bulbitech.com/.

10. Mahanama B, Jayawardana Y, Rengarajan S, Jayawardena G, Chukoskie L, Snider J, et al. Eye Movement and Pupil Measures: A Review. Frontiers in Computer Science 2022;3.

11. Pupil Labs. Pupil Labs Neon. 2024; Available at: https://pupil-labs.com/products/neon.

12. Holmqvist K. Eye tracking : a comprehensive guide to methods and measures. Oxford: Oxford University Press; 2011.

13. Blekher T, Weaver MR, Cai X, Hui S, Marshall J, Jackson JG, et al. Test-retest reliability of saccadic measures in subjects at risk for Huntington disease. Invest Ophthalmol Vis Sci 2009 Dec;50(12):5707–5711.

14. Weir JP. Quantifying test-retest reliability using the intraclass correlation coefficient and the SEM. J Strength Cond Res 2005 Feb;19(1):231–240.

15. Nij Bijvank JA, Petzold A, Balk LJ, Tan HS, Uitdehaag BMJ, Theodorou M, et al. A standardized protocol for quantification of saccadic eye movements: DEMoNS. PLoS One 2018 Jul 16;13(7):e0200695.

16. Martin Bland J, Altman D. STATISTICAL METHODS FOR ASSESSING AGREEMENT BETWEEN TWO METHODS OF CLINICAL MEASUREMENT. The Lancet 1986;327(8476):307–310.

17. Martins AJ, Kowler E, Palmer C. Smooth pursuit of small-amplitude sinusoidal motion. J Opt Soc Am A 1985 Feb;2(2):234–242.

18. Salman MS, Sharpe JA, Lillakas L, Dennis M, Steinbach MJ. Smooth pursuit eye movements in children. Exp Brain Res 2006 Feb;169(1):139–143.

19. Schröder R, Keidel K, Trautner P, Radbruch A, Ettinger U. Neural mechanisms of background and velocity effects in smooth pursuit eye movements. Hum Brain Mapp 2023 Feb 15;44(3):1002–1018.

20. Gersch TM, Kowler E, Schnitzer BS, Dosher BA. Attention during sequences of saccades along marked and memorized paths. Vision Res 2009 Jun;49(10):1256–1266.

21. Hutton SB. Cognitive control of saccadic eye movements. Brain Cogn 2008 Dec;68(3):327–340.

22. Pierce, J.E., Clementz, B.A., McDowell, J.E. Saccades: Fundamentals and Neural Mechanisms. In: Klein, C., Ettinger, U, editor. Eye Movement Research: Springer, Cham; 2019.

23. Mathôt S. Pupillometry: Psychology, Physiology, and Function. J Cogn 2018 Feb 21;1(1):16.

24. Wickremasinghe SS, Smith GT, Stevens JD. Comparison of dynamic digital pupillometry and static measurements of pupil size in determining scotopic pupil size before refractive surgery. J Cataract Refract Surg 2005 Jun;31(6):1171–1176.

25. Schmitz S, Krummenauer F, Henn S, Dick HB. Comparison of three different technologies for pupil diameter measurement. Graefes Arch Clin Exp Ophthalmol 2003 Jun;241(6):472–477.

